# Influence of ethnicity and consanguinity on the genetic architecture of Hypertrophic Cardiomyopathy: insights from an understudied population

**DOI:** 10.1101/2022.10.09.22280408

**Authors:** Mona Allouba, Roddy Walsh, Alaa Afify, Mohammed Hosny, Sarah Halawa, Aya Galal, Mariam Fathy, Pantazis I. Theotokis, Ahmed Boraey, Amany Ellithy, Rachel Buchan, Risha Govind, Nicola Whiffin, Shehab Anwer, Ahmed ElGuindy, James S. Ware, Paul J.R. Barton, Yasmine Aguib, Magdi Yacoub

## Abstract

Hypertrophic cardiomyopathy (HCM) is an inherited cardiac condition characterized by phenotypic heterogeneity that could partly be explained by the variability in genetic variants contributing to disease. Accurate interpretation of these variants constitutes a major challenge for diagnosis and implementing precision medicine, especially in understudied populations. Here, we leverage ancestry-matched Egyptian patients (n=514) and deeply-phenotyped controls (n=400) to accurately define the genetic architecture of HCM. We also compare HCM variation between Egyptian and predominantly European patients to identify genetic features unique to consanguineous populations in Middle East and North Africa (MENA), which are likely to represent important contributors to disease. We report a higher prevalence of homozygous variants in Egyptian patients (4.1% vs 0.1%, p-value=2×10^×7^), with variants in the minor HCM genes *MYL2, MYL3* and *CSRP3* more likely to present in homozygosity than the major genes (*MYH7, MYBPC3*), suggesting that these variants are less penetrant in heterozygosity. Biallelic variants in the recessive HCM gene *TRIM63* were detected in 2.1% of patients (5-fold greater than European patients), highlighting the importance of recessive inheritance/genes in consanguineous populations. Finally, significantly fewer rare variants detected in Egyptian HCM patients could be classified as (likely) pathogenic compared to Europeans (40.8% vs. 61.6%, p-value=1.6×10^×5^) due to the underrepresentation of MENA populations in current HCM databases. This proportion increased to 53.8% after incorporating methods that compare variant frequencies between Egyptian patients and controls. Together, our findings demonstrate that studying consanguineous populations reveals novel insights with relevance to genetic testing and our understanding of the genetic architecture of HCM.

## Introduction

Hypertrophic cardiomyopathy (HCM [MIM: 192600] is a common inherited cardiac condition affecting at least 1:500 in the general population ^1,2^. It is defined by left ventricular hypertrophy (LVH) in the absence of other identifiable cardiac or systemic causes. Since its first description in 1958, HCM has been considered as a highly heterogenous disease with regards to mode of inheritance, phenotype and genotype ^3–5^. The genetic etiology of HCM is largely attributed to variation in genes that encode components of the contractile apparatus (sarcomere) in cardiac muscle cells.

Large HCM datasets from the Sarcomeric Human Cardiomyopathy Registry (SHaRe) and other diverse ancestral HCM populations have provided strong evidence supporting disease-associated, long-term adverse clinical outcomes, including mortality ^6–11^. With respect to genotype, SHaRe demonstrated that patients with pathogenic sarcomere variants had an earlier onset of disease and were at a higher risk of developing adverse clinical outcomes ^10^. Given the interplay between genetics and disease progression, these patients may benefit from effective treatments that delay or prevent HCM-related complications provided these pathogenic variants are well characterized ^10,12^. Although immense progress has been made in elucidating the genetic determinants of HCM, much remains to be explored including genetic-, environmental- and epigenetic factors influencing disease penetrance and expressivity. Also, the rapid integration of genetic testing into everyday clinical practice has resulted in the identification of a large number of variants of uncertain significance, which cannot be acted upon clinically^10,13,14^. This issue is more evident in underrepresented populations, who are more likely to receive inconclusive genetic testing results, compared to well-studied European-ancestry HCM patients ^15^. Therefore, extending genomic research to diverse ancestral populations will not only reduce health disparities as we move towards precision-based HCM patient management but may also enhance variant interpretation for diagnosis, and provide novel genetic and mechanistic insights into the disease. For example, we have recently reported the identification of a new class of HCM pathogenic variants in *MYH7*, which was enriched in Egyptian patients over ancestry-matched controls and absent from large-scale HCM patients of predominantly European ancestry ^16–18^.

The aim of this study is to define the genetic architecture of HCM in the previously understudied Egyptian population that is known to have a high rate of consanguinity ^19,20^. This study provides, to our knowledge, the largest and most comprehensive analysis of HCM in the Middle East and North Africa (MENA) region to date. We leverage a prospective Egyptian HCM cohort and an ancestry-matched, Egyptian healthy volunteer control cohort with comprehensive cardiovascular phenotyping, to define the genetic architecture of HCM in Egypt. In addition, we compare genetic data between Egyptian and well-characterized patients of predominantly European ancestry to identify genetic features unique to consanguineous populations of MENA ancestry which are likely to be important contributors of HCM.

## Methods

### Study population

#### Egypt HCM cohort

A prospective cohort of unrelated HCM patients (n=514) was recruited to the Aswan Heart Centre (AHC), Egypt as part of the Egyptian Collaborative Cardiac Genomics (ECCO-GEN) Project and the National HCM Registry. This cohort is hereafter referred to as Egypt HCM. Clinical diagnosis of HCM was confirmed by echocardiography and/or Cardiac Magnetic Resonance (CMR) imaging.

#### UK HCM cohort

A prospective cohort of HCM patients of predominantly European ancestry (n=684) was recruited by the NIHR Cardiovascular Biomedical Research Unit at the Royal Brompton Hospital (RBH), London ^21^. This cohort is hereafter referred to as UK HCM. HCM diagnosis was confirmed with CMR.

#### Egypt control cohort

Egyptian healthy volunteers (n=400) were recruited from across the country as part of the Egyptian Collaborative Cardiac Genomics (ECCO-GEN) Project aiming to study local genetic variation in 1000 healthy individuals ^17^. All volunteers underwent detailed cardiovascular phenotyping, including clinical examination, 12-lead electrocardiogram, and CMR to confirm absence of CVD. This cohort is hereafter referred to as Egypt control.

#### UK control cohort

UK healthy volunteers (n=1,054) of predominantly European ancestry were recruited prospectively via advertisement for the UK Digital Heart Project at Imperial College London ^22^. All volunteers, denoted hereafter as UK controls, underwent CMR to confirm the absence of cardiac disease.

Participants from all four cohorts provided informed consent and were approved by their local research ethics committees.

#### Gene selection

DNA samples from the above-mentioned case and control cohorts were sequenced on Illumina platforms (Miseq, Nextseq or Hiseq) and a subset of these on Life Technologies SOLiD 5500xl using cardiac gene panels including the TruSight Cardio Sequencing kit ^23^. The number of HCM patients and controls from the Egypt and UK cohorts analyzed for each gene are depicted in Supplementary Table 1. Thirteen validated autosomal dominant HCM genes, including sarcomere-encoding (*ACTC1* (MIM: 102540), *MYBPC3* (MIM: 600958), *MYH7* (MIM:160760*), MYL2* (MIM: 160781), *MYL3* (MIM: 160790), *TNNC1* (MIM: 191040), *TNNI3* (MIM: 191044), *TNNT2* (MIM: 191045) and *TPM1* (MIM: 191010)) and non-sarcomere-encoding genes (*PLN* (MIM: 172405), *ACTN2* (MIM: 102573), *CSRP3* (MIM: 600824), *JPH2* (MIM: 605267)) were analyzed ^21,24,25^. Genes, which have been previously shown to account for less than 5% of HCM cases were defined in this work as “minor” genes ^18^. *MYH7* and *MYBPC3* were defined here as “major” genes as they account for over 70% of identified pathogenic variants in HCM ^26^. Separately, biallelic variants in the recently implicated recessive HCM gene *TRIM63* (MIM: 606131) were evaluated. Some genes recently identified as rarer causes of HCM (*ALPK3* (MIM: 617608), *FHOD3* (MIM: 609691, *FLNC* (MIM: 102565) were not included on the gene panels used in this study.

#### Rare variant filtering and classification

Rare variants in validated HCM genes were defined as having a filtering allele frequency (FAF) of ≤ 4×10^×5^ in gnomAD (v2.1.1) based on the statistical framework proposed by Whiffin *et al*., 2017 ^27^. Variants were annotated by CardioClassifier based on rules defined by the American College of Medical Genetics and Genomics/Association of Molecular Pathology (ACMG-AMP) as Pathogenic (P), Likely Pathogenic (LP) and Uncertain Significance (VUS) (full details in Supplementary Methods) ^28,29^. Detailed lists of all rare variants observed in the Egyptian and UK HCM and control cohorts are provided in Supplementary Tables 2-5.

#### Comparison of the genetic landscape of HCM between Egypt and UK HCM cohorts

First, the genetic architecture of the HCM in the Egyptian population was assessed by comparing the frequency of rare variation in validated HCM genes between ancestry-matched patients and controls. Second, the genetic landscape of HCM was compared between the Egypt and UK HCM cohorts using their respective population-matched controls. For each gene, the excess of rare variants in the Egypt HCM cohort over the Egypt control cohort was compared with the excess of rare variants in the UK cohort over UK controls (Egypt HCM_Control_ excess vs. UK HCM_Control_ excess). Third, the proportion of genotype-positive patients (i.e., those carrying rare variants) attributable to each HCM gene was compared between the Egypt and UK HCM cohorts.

#### Comparison of the proportion of clinically actionable variants between the Egypt and UK HCM cohorts

The proportion of clinically actionable variants (i.e., P and LP) was evaluated in the Egypt HCM cohort by dividing the number of patients with P/LP variants by the total number of patients with rare variants in validated HCM genes. For patients with multiple hits (refer to Supplementary Tables 6 and 7), the variant with the highest pathogenicity was prioritized (i.e., P then LP then VUS). The proportion of clinically actionable variants was then compared between the Egypt and UK HCM cohorts.

#### Application of the new Etiological Fraction (EF)-based American College of Medical Genetics and Genomics/Association or Molecular Pathology (ACMG-AMP) rule on rare *MYH7* missense variants

The effect of the recently proposed EF-based ACMG/AMP rules by Walsh *et al*., 2019 on the yield of actionable *MYH7* missense variants (i.e. P and LP) identified in the Egypt HCM cohort was assessed^13^.

First, we evaluated whether the regional distributions of pathogenic and benign variation in *MYH7*, previously observed in European individuals, is also observed in Egyptians. The frequency of rare missense *MYH7* variants in the predefined patient-enriched HCM cluster (residues: 167-931) was calculated ^13^. The distribution of rare missense *MYH7* variants in the Egypt HCM and control cohorts and was visualized using the TrackViewer package (v 1.22.1) in R statistical environment (v 3.6.3). This was further compared with the previously published cohort of >6,000 HCM patients from Partners Laboratory of Molecular Medicine and Oxford Medical Genetics Laboratory (denoted here as LMM/OMGL) ^18^. Second, the odds ratio (OR) and etiological fraction (EF) (EF = (OR-1)/OR) for this cluster were calculated using the Egypt control cohort as the reference control population.

*MYH7* missense variants were classified as per the adopted new EF-based PM1_strong rule, which is activated if these variants are located in a protein region with EF≥0.95

## Results

### Cohort characteristics

The Egypt HCM cohort comprised 514 patients (67% male, 33% female; full details in Supplementary Table 8). Consanguinity was reported in 28.4% of Egyptian patients. Notably, UK patients (n=684) were ca. 20 years older (mean onset age 55.5±15.5 years vs. 34.7 ± 17.4 years, p-value≤2.2×10^×16^), presented with less severe hypertrophy (LVMWT: 18.9±4.4mm vs. 23.6±7mm, p-value=4.9×10^×15^) and showed a lower incidence of left ventricular outflow tract obstruction (LVOTO) (24%) compared to Egyptian patients (36% vs. 73%, p-value≤2.2×10^×16^). A summary of the demographic- and cardiac characteristics of the Egypt and UK HCM cohorts with available data is outlined in Table 1.

**Table 1:** Demographic and cardiac characteristics of Egypt and UK HCM cohorts.

| General characteristics | Egypt HCM n(%) | UK HCM n(%) | p-value |
| --- | --- | --- | --- |
| Demographics |  |  |  |
| Age (years) (mean (sd)) | $34.7 \pm 17.4$ | $55.5 \pm 15.5$ | $< 2.2 \times 10^{-16}$ |
| Male gender n(%) | 345 (67.3%) | 473 (73.4%) | 0.02 |
| Offspring of Consanguineous Marriage* | 99 (28.4%) | N/A | N/A |
| FH of HCM | 111 (25.1%) | 127 (24.5%) | 0.88 |
| FH of SCD | 78 (17.6%) | 116 (25.2%) | 0.01 |
| Cardiac Characteristics |  |  |  |
| LVMWT (mm)** (mean (sd)) | $23.6 \pm 7$ | $18.9 \pm 4.4$ | $4.9 \times 10^{-15}$ |
| Extreme LVMWT (>30mm) | 54 (17.5%) | 8 (1.3%) | $< 2.2 \times 10^{-16}$ |
| LVOTO ($\geq 30$ mm Hg) | 220 (72.8%) | 223 (36.1%) | $< 2.2 \times 10^{-16}$ |
| LVEF (%)** (mean (sd)) | $71.5 \pm 13.3$ | $74.3 \pm 8.5$ | 0.004 |
| Myectomy | 168 (51.5%) | N/A | N/A |
| ICD implantation | 7 (2.7%) | N/A | N/A |

\*Consanguinity (self-reported) was defined as first-cousin marriage. FH: Family History, SCD: Sudden Cardiac Death, LVMWT: Left Ventricular Maximal Wall Thickness, LVOTO: Left Ventricular Outflow Tract Obstruction, LVEF: Left Ventricular Ejection Fraction, ICD: Implantable Cardioverter Defibrillator. \*\*LVMWT and LVEF were measured in the Egypt HCM and UK HCM cohorts by echocardiography and CMR, respectively.

### Differences in the genetic architecture of HCM between Egyptian- and UK patients

In order to evaluate the genetic architecture of HCM in Egypt, we compared the burden of rare variation (gnomAD_popmax_ FAF ≤4×10^×5^) in validated HCM genes between Egyptian patients and controls (Figure 1A, left). The inclusion of ancestry-matched controls into this analysis enabled us to address the potential bias introduced by defining rare variants using gnomAD, which has minimal representation from the MENA region (Supplementary Figure 1).

**Figure 1:**
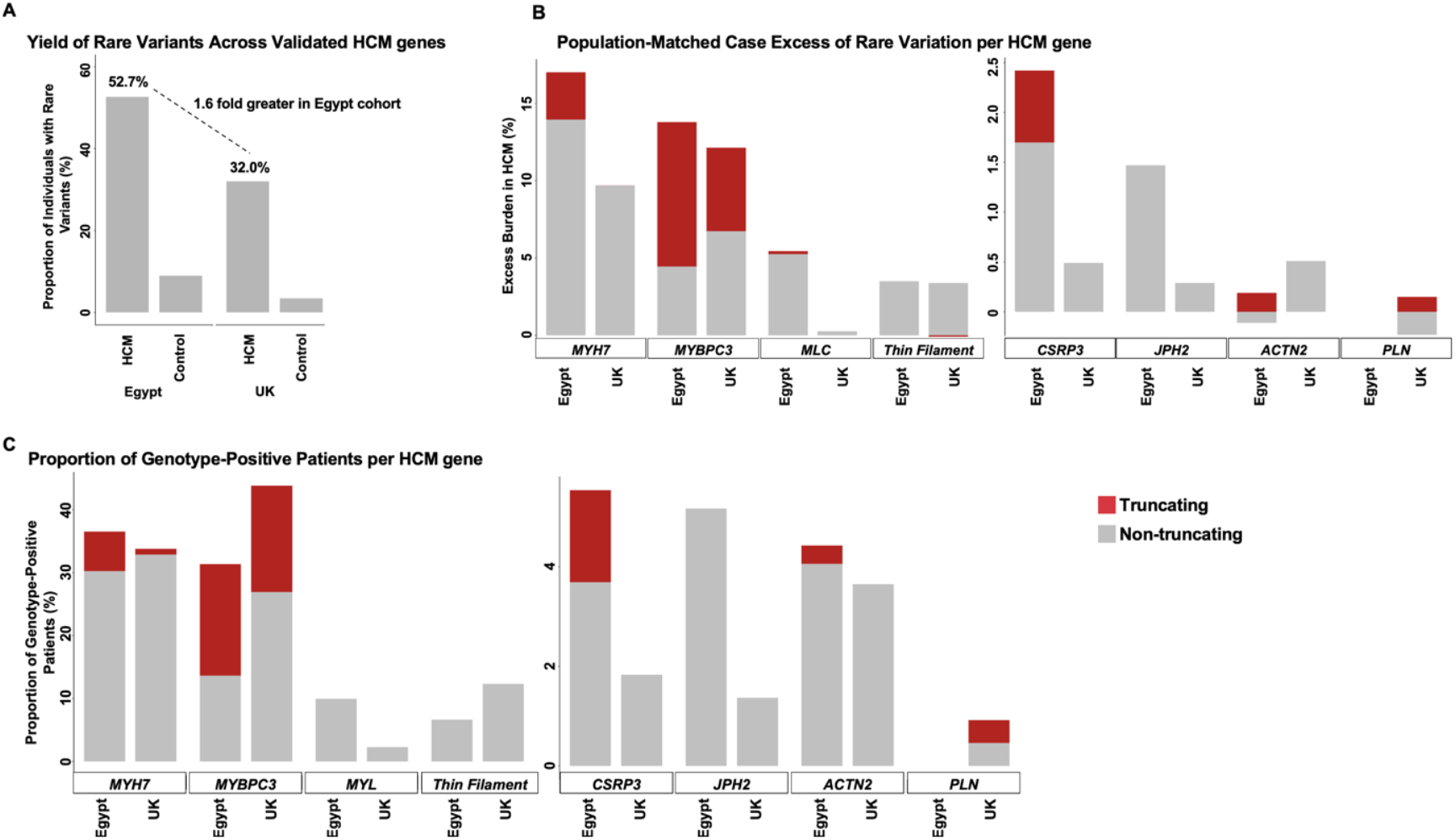
Enrichment of rare variation in HCM minor genes (MLC, *CSRP3 & JPH2*) in the Egypt HCM cohort vs. UK HCM. **A)** The overall frequency of rare variants (gnomAD_popmax_ FAF ≤4×10^×5^) in 13 validated HCM genes was compared between Egypt and UK HCM patients and controls. A 1.6 fold greater excess burden (i.e. total case frequency – total control frequency) was observed in the Egypt HCM cohort, which should be accounted for with the direct gene comparisons (B). **B)** Population-matched patients vs. control comparison (Egypt HCM_Control_ excess and UK HCM_Control_ excess) of genetic variation between Egypt and UK cohorts for each HCM gene/gene class. C) Comparison of the proportion of genotype-positive patients (i.e., those carrying rare variants) per HCM gene confirms the enrichment of rare variants in the HCM minor genes MLC (*MYL2* & *MYL3*), *CSRP3* and *JPH2* in Egyptians (as observed in Figure B). Truncating and non-truncating variants (Figures A & B) are shown in red and grey, respectively. *Thin filament: ACTC1, TNNC1, TNNI3, TNNT2, TPM1*.

In addition, we compared the genetic landscape of HCM between the Egypt and UK HCM cohorts (using their respective population-matched controls), to directly compare the excess burden of rare variants in HCM-associated genes in each cohort (labelled as Egypt HCM_Control_ excess and UK HCM_Control_ excess) (Figure 1A, right). A higher overall excess burden of rare variation was observed in the Egypt HCM cohort compared to UK HCM (52.7% vs. 32.0%) (Figure 1A), with 1.6-fold more Egyptian patients carrying rare variants compared to UK HCM (Figure 1B). We then performed a secondary analysis comparing the proportion of genotype-positive patients (i.e., those carrying rare variants) attributable to each HCM gene between the Egypt and UK HCM cohorts (Figure 1C).

As expected, rare variants in both *MYH7* and *MYBPC3* accounted for the majority of rare variation in both the Egypt and UK HCM cohorts (Table 2). *MYBPC3* non-truncating variants and *PLN* variants accounted for a higher proportion of genotype-positive patients in the UK HCM cohort compared to Egyptian patients (*MYBPC3*: 26.9% vs. 13.7% and *PLN*: 0.92% vs. 0%) (Table 2). In contrast, *MYH7* truncating variants accounted for a higher proportion of genotype-positive patients in the Egypt HCM cohort, which is explained by the recurrent variant c.5769delG [p.Ser1924AlafsTer9]. This variant was present in 3.3% of Egypt HCM patients and we recently showed that it causes HCM by introducing a distal premature termination codon, which escapes nonsense mediated decay resulting in the expression of an abnormal protein^16^. Several recurrent truncating variants in *MYBPC3* were also detected, with two of these (c.1516delG [p.Asp506ThrfsTer7] (0.97%, n=5) and c.534_541delGGCCGGCG [p.Ala179GlnfsTer59] (1.36%, n=7)) likely to be Egyptian/North African specific founder variants. *MYBPC3* founder variants have similarly been reported in numerous regions ^30^.

**Table 2:** Comparison of population-matched patient versus control excess frequencies (left) and the proportion of genotype-positive patients between Egypt HCM and UK HCM cohorts (right) for sarcomeric- and other validated HCM genes.

| Gene/Gene groups | Variant type | Excess of RV in patients vs. controls |  | Proportion of all G+ |  |
| --- | --- | --- | --- | --- | --- |
|  |  | Egypt HCM<sub>Control</sub> excess | UK HCM<sub>Control</sub> excess | Egypt HCM G+ prop | UK HCM G+ prop |
| *MYH7* | Non-truncating | 13.95% | 9.68% | 30.26% | 32.88% |
|  | Truncating | 3.06% | 0.01% | 6.27% | 0.91% |
| *MYBPC3* | Non-truncating | 4.45% | 6.73% | 13.65% | 26.94% |
|  | Truncating | 9.34% | 5.41% | 17.71% | 16.89% |
| *Myosin light chain (MLC)* | Non-truncating | 5.25% | 0.26% | 9.96% | 2.28% |
|  | Truncating | 0.19% | 0.00% | 0.37% | 0% |
| *Thin Filament* | Non-truncating | 3.50% | 3.38% | 6.64% | 12.33% |
|  | Truncating | 0.00% | -0.09% | 0% | 0% |
| *CSRP3* | Non-truncating | 1.70% | 0.49% | 3.69% | 1.83% |
|  | Truncating | 0.72% | 0.00% | 1.85% | 0% |
| *JPH2* | Non-truncating | 1.47% | 0.29% | 5.17% | 1.37% |
|  | Truncating | 0% | 0% | 0% | 0% |
| *ACTN2* | Non-truncating | -0.11% | 0.51% | 4.06% | 3.65% |
|  | Truncating | 0.19% | 0% | 0.37% | 0% |
| *PLN* | Non-truncating | 0% | -0.23% | 0% | 0.46% |
|  | Truncating | 0% | 0.15% | 0% | 0.46% |

Due to the limited number of observed variants, HCM genes encoding components of the thin filament (*ACTC1, TNNC1, TNNI3, TNNT2* and *TPM1*) and myosin light chain (MLC: *MYL2* and *MYL3*) were combined into groups based on their similar function. While rare variation in thin filament genes accounted for a relatively higher proportion of genotype-positive patients in the UK HCM cohort compared to Egypt HCM (12.3% vs. 6.6%), non-truncating variants in MLC genes accounted for a higher proportion of genotype-positive patients in the Egypt HCM cohort (9.96% vs. 2.28%) (Table 2).

Six distinct variants were identified in each of *MYL2* (5 missense and 1 splice site) and *MYL3* (6 missense) (Supplementary Table 2). Of these, three *MYL2* variants were recurrently observed in Egyptian patients (c.173G>A [p.Arg58Gln] (n=2,0.39%), c.450G>T [p.Leu150Phe] (n=3,0.58%) and c.278C>T [p.Ala93Val] (n=5, 0.97%)). In *MYL3* 3/6 missense variants observed in Egyptian patients were also recurrent (c.460C>T [p.Arg154Cys] (n=2,0.39%), c.518T>C [p.Met173Thr] (n=4,0.78%) and c.508G>C [p.Glu170Gln] (n=6,1.17%)). With the exception of the known pathogenic *MYL2* p.Arg58Gln variant, these recurrent variants were absent from both the UK HCM cohort and LMM/OMGL HCM cohorts. Also, the *MYL2* variant p.Leu150Phe has not been previously reported whereas the other ten MLC variants are currently classified as VUS in ClinVar.

For the non-sarcomeric HCM genes, rare variants in both *CSRP3* (truncating and non-truncating) and *JPH2* (non-truncating) showed a nominal enrichment in the genotype-positive Egypt HCM cohort compared to UK HCM (Table 2).

To explore why variants in some minor HCM genes (MLC, *CSRP3* and *JPH2*) seem to be more prevalent in the Egypt HCM cohort compared to UK HCM, we then examined the effect of consanguinity, and the frequency of homozygous variants, on this cohort.

### Higher burden of homozygous variants in the Egypt HCM cohort compared to UK HCM

In the Egypt HCM cohort, 28% of patients reported to have consanguineous parents. This prompted us to evaluate the frequency of homozygous variants, both for rare variants (FAF_popmax_≤4.0×10^×5^) and for low frequency variants sufficiently rare to cause biallelic disease (FAF_popmax_≤0.00126) ^27^. The yield of homozygous variants was then compared with the UK cohort.

As expected, homozygous variants were significantly more common in Egyptian patients compared to UK, for both the rare (4.1% (n=21) vs. 0.1% (n=1), p=2×10^×7^) and low frequency (2.1% (n=11) vs. 0%, p=8.5×10^×5^) variants. Table 3 summarizes the homozygous variants observed in Egyptian patients. Homozygous variants in these genes were not observed in either the Egypt or UK control cohorts. Among Egypt HCM patients carrying homozygous variants, 31.3% reported not having consanguineous parents (i.e. first cousins), which may indicate an underreporting of consanguinity or more distantly related patients.

**Table 3:** Details of homozygous variants identified in the Egypt HCM cohort (n=514) compared to Egypt controls (n=400)

| Gene | Variant type | cDNA variant | Protein variant | gnomAD<br>FAF<sub>popmax</sub> | Control hom<br>freq (count) | Control<br>het freq<br>(count) | HCM hom<br>case freq<br>(count) | HCM het<br>case freq<br>(count) |
| --- | --- | --- | --- | --- | --- | --- | --- | --- |
| **A) Sufficiently rare variants (gnomAD FAF<sub>popmax</sub> ≤ 0.00126) to cause mono allelic HCM** |  |  |  |  |  |  |  |  |
| MYH7 | Missense | c.5122G>A | p.Glu1708Lys | 0 | 0% | 0% | 0.19% (1) | 0% |
|  | Missense | c.4258C>T | p.Arg1420Trp | 3.0x10<sup>-6</sup> | 0% | 0% | 0.19% (1) | 0.39% (2) |
|  | Missense | c.3622G>C | p.Asp1208His | 0 | 0% | 0% | 0.19% (1) | 0.58% (3) |
|  | Missense | c.1064C>T | p.Ala355Val | 0 | 0% | 0% | 0.19% (1) | 0% |
|  | Missense | c.925G>A | p.Asp309Asn | 1.1x10<sup>-5</sup> | 0% | 0% | 0.19% (1) | 0.78% (4) |
|  | Frameshift* | c.5769delG | p.Ser1924AlafsTer9 | 0 | 0% | 0% | 0.19% (1) | 3.11% (16) |
| MYBPC3 | Missense | c.2908C>T | p.Arg970Trp | 0 | 0% | 0.25% (1) | 0.19% (1) | 0.39% (2) |
|  | Missense | c.2458C>G | p.Arg820Gly | 0 | 0% | 0% | 0.19% (1) | 0% |
|  | Inframe<br>Deletion | c.2441_2443delA<br>GA | p.Lys814del | 0 | 0% | 0% | 0.19% (1) | 0% |
|  | Nonsense | c.1558G>T | p.Glu520Ter | 0 | 0% | 0% | 0.19% (1) | 0% |
|  | Splice acceptor | c.1227-2A>G | - | 0 | 0% | 0% | 0.19% (1) | 0.19% (1) |
| MYL2 | Missense | c.278C>T | p.Ala93Val | 0 | 0% | 0% | 0.19% (1) | 0.78% (4) |
| MYL3 | Missense | c.508G>C | p.Glu170Gln | 0 | 0% | 0% | 0.58% (3) | 0.58% (3) |
| TNNI3 | Missense | c.485G>T | p.Arg162Leu | 0 | 0% | 0% | 0.19% (1) | 0% |
| CSRP3 | Splice acceptor | c.415-1G>C | - | 0 | 0% | 0% | 0.19% (1) | 0% |
|  | Splice donor | c.414+1G>T | - | 0 | 0% | 0.25% (1) | 0.58% (3) | 0.19% (1) |
|  | Missense | c.365G>A | p.Arg122Gln | 2.9x10<sup>-6</sup> | 0% | 0% | 0.19% (1) | 0.58% (3) |
| **B) Sufficiently rare variants (gnomAD FAF<sub>popmax</sub> ≤ 0.00126) to cause biallelic HCM** |  |  |  |  |  |  |  |  |
| MYBPC3 | Missense | c.3676C>T | p.Arg1226Cys | 6.4x10<sup>-5</sup> | 0% | 0% | 0.19% (1) | 0% |
|  | Missense | c.2618C>A | p.Pro873His | 9.3x10<sup>-5</sup> | 0% | 0% | 0.19% (1) | 0% |
|  | Missense | c.1321G>A | p.Glu441Lys | 2.0x10<sup>-4</sup> | 0% | 2.0% (8) | 0.39% (2) | 4.3% (22) |
| MYL3 | Missense | c.530A>G | p.Glu177Gly | 4.4x10<sup>-5</sup> | 0% | 0.75% (3) | 0.78% (4) | 0.19% (1) |
|  | Missense | c.170C>A | p.Ala57Asp | 4.0x10<sup>-4</sup> | 0% | 0% | 0.39% (2) | 0.19% (1) |
| JPH2 | Missense | c.572C>G | p.Pro191Arg<sup>‡</sup> | 2.0x10<sup>-4</sup> | 0% | 0.25% (1) | 0.19% (1) | 0.19% (1) |

A) Summary of Egyptian variants that are sufficiently rare (gnomAD FAF<sub>popmax</sub> ≤ 4x10<sup>-5</sup>) to cause mono allelic disease. B) Additional variants that are sufficiently rare (gnomAD FAF<sub>popmax</sub> ≤ 0.00126) to cause biallelic disease are also shown. \*The MYH7 variant c.5769delG was caused by a nucleotide deletion that was predicted to ultimately give rise to a near full-length protein (i.e., only 4 AAs shorter than the wildtype). The control frequency represents heterozygous counts (no homozygous variants were identified). <sup>‡</sup>This variant is unlikely to be pathogenic as it co-occurred with the pathogenic MYH7 variant p.Arg870His in the HCM patient (also, it's allele frequency in gnomAD-Ashkenazi Jewish population is 0.007215).

Two Egyptian patients presented with homozygous *MYBPC3* truncating variants (c.1558G>T [p.Glu520Ter] and c.1227-2A>G). Truncating variants in *MYBPC3* are known to lead to severe, early-onset disease when expressed in homozygosity ^31^. The patient carrying the nonsense variant p.Glu520Ter was indeed diagnosed with HCM at birth and died suddenly before adolescence. In contrast, the patient carrying the c.1227-2A>G splice variant in homozygosity presented with HCM in adulthood, although at a younger age and with more severe hypertrophy (LVMWT: 27mm vs. 20mm) than the patient with the same variant in the heterozygous state. Accordingly, this splice acceptor variant is predicted (by SpliceAI) to result in an exon extension of 33 intronic bases into the *MYBPC3* transcript, which suggests that the variant maintains the open reading frame and may therefore function as an inframe insertion (Supplementary Figure 2).

Notably, homozygous variants were particularly enriched in three minor HCM genes (i.e. those previously shown to account for less than 5% of HCM patients each in European populations) - *CSRP3* and the MLC genes *MYL2* and *MYL3* ^18^ (Table 3). Therefore, we sought to compare the relative contribution of homozygous variants across the HCM genes. The proportion of rare homozygous variants was consistent across the major HCM genes, with 5-7% of variants being homozygous in *MYH7, MYBPC3* and the thin filament genes (Table 4 and Figure 2). This proportion could be regarded as a background homozygosity rate for dominant HCM genes caused by the level of consanguinity present in the Egyptian population. In contrast the MLC and *CSRP3* genes showed a substantially higher proportion of homozygous variants (25.6% and 33.3%, respectively). It is worth mentioning that the increase in frequency of MLC homozygous variants from 14.3% (using the rare frequency threshold) to 25.6% (using the biallelic frequency threshold) was largely due to a recurrently observed low-frequency (gnomAD_popmax_ FAF=4.4×10^×5^) *MYL3* variant c.530A>G [p.Glu177Gly] in the Egyptian cohort (0.78%,n=4).

**Table 4:** Relative contribution of homozygous variants for validated autosomal dominant HCM genes.

| Gene/Gene groups | Variants with gnomAD $FAF_{popmax} \leq 4 \times 10^{-5}$ | | Additional variants with gnomAD $FAF_{popmax} \leq 0.00126$ | | Total hom freq (count/total) |
| --- | --- | --- | --- | --- | --- |
|  | Total no. of patients | Hom case freq(count) | Total no. of patients | Hom case freq(count) |  |
| *MYH7* | 99 | 6.1% (6) | 5 | 0% | 5.8% (6/104) |
| *MYBPC3* | 85 | 5.9% (5) | 45 | 8.9% (4) | 6.9% (9/130) |
| *CSRP3* | 15 | 33.3% (5) | 0 | 0% | 33.3% (5/15) |
| *Myosin light chain* | 28 | 14.3% (4) | 11 | 54.5% (6) | 25.6% (10/39) |
| *Thin filament* | 18 | 5.6% (1) | 8 | 0% | 3.8% (1/26) |
| *ACTN2* | 12 | 0% | 8 | 0% | 0% (0/20) |
| *JPH2* | 14 | 0% | 4 | 25% (1) | 5.6% (1/18) |
| *PLN* | 0 | 0% | 0 | 0% | 0% |

**Figure 2:**
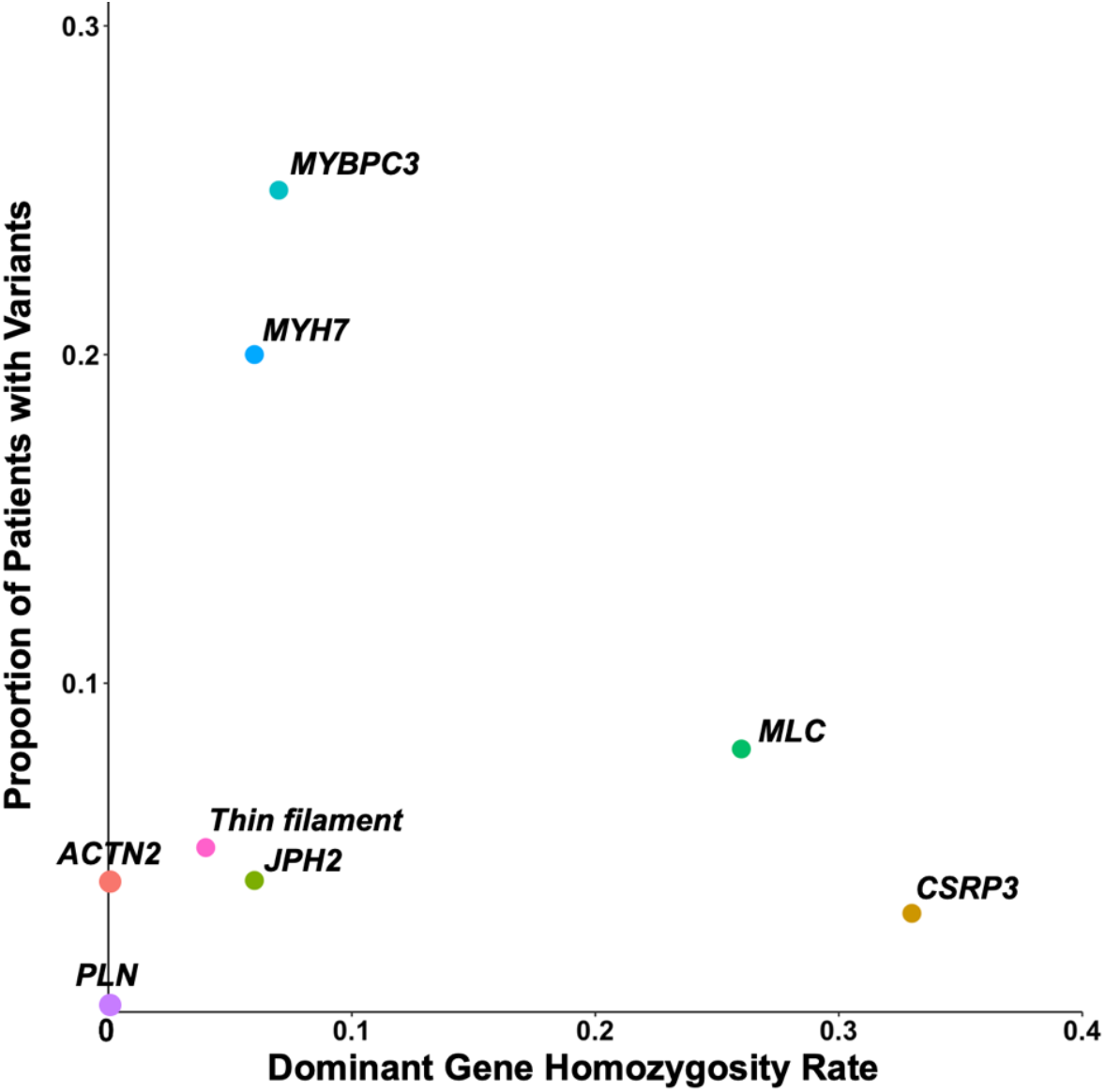
Comparison of the rate of homozygosity across the analyzed dominant HCM genes. MLC- (*MYL2, MYL3*) and *CSRP3* genes showed a markedly higher rate of homozygosity compared to major HCM genes (*MYBPC3, MYH7*) and thin filament genes (*ACTC1, TNNC1, TNNI3, TNNT2, TPM1*).

These findings suggest that, unlike the major HCM genes, rare variants in these minor genes may be generally less penetrant in the heterozygous state. Evidence for an additive, rather than a true recessive, model is supported by the fact that heterozygous variants in these genes are also enriched in Egyptian patients compared to controls and that the vast majority of previously reported pathogenic CSRP3 and missense MLC variants have been associated with autosomal dominant HCM. Taken together, these findings explain the relatively higher proportion of rare variants in MLC and CSRP3 genes observed in Egyptian patients compared to UK (Figure 1), with high consanguinity rates enabling variants to occur in a more penetrant homozygous state.

### A significantly higher proportion of VUSs is observed in the Egypt HCM cohort compared to UK HCM

In contrast to MENA populations, European HCM patients have been extensively studied over the past decades. As a result, they are more likely to carry a disease-causing variant that has been previously well-characterized. Therefore, we hypothesize that in patients with rare variants in validated HCM genes, Egyptians have a higher proportion of VUSs compared to the UK HCM cohort.

The proportion of patients with rare variants in HCM genes was 46.7% (n=240) and 28.9% (n=198) in the Egypt and UK HCM cohorts, respectively. As expected, the proportion of VUSs was significantly higher in the Egypt HCM cohort compared to UK (59.2% vs. 38.4%, p-value=1.6×10^×5^), which highlights the challenge in interpreting variants in understudied populations (Figure 3).

**Figure 3:**
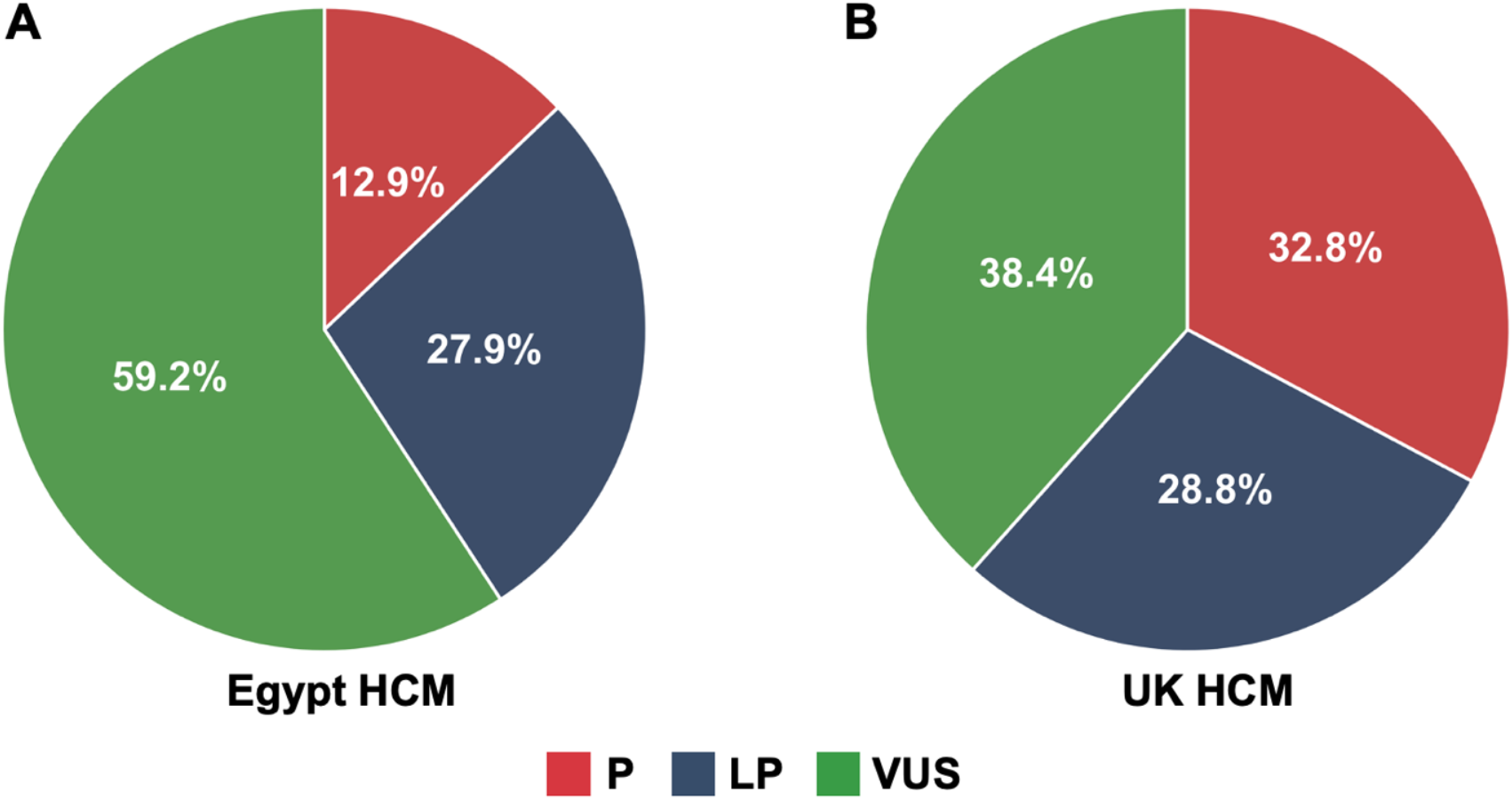
Higher proportion of Variants of Uncertain Significance (VUSs) is observed in (A) Egyptian patients compared to (B) UK. For patients with rare variants in validated HCM genes, the distribution of rare variants by variant classification (pathogenic (P), Likely Pathogenic (LP) and VUSs) is shown. For patients with multiple hits (Supplementary Tables 6 and 7) the variant with the highest pathogenicity was prioritized (i.e., P then LP then VUS). The proportion of VUSs was significantly higher in Egypt HCM patients vs. UK (59.2% vs. 38.4%, p-value=1.6×10^×5^).

### Analysis of HCM rare variation in the ancestry-matched Egyptian case-control cohort increases the proportion of clinically actionable variants

The availability of genetic data from a deeply-phenotyped control cohort of 400 Egyptians as well as cohort-level analysis of the HCM patients described here enabled us to re-evaluate the classification of identified VUSs in patients by adopting 3 approaches. First, we examined whether these VUSs were prevalent among our control cohort (i.e. n≥2; MAF≥0.0025), which would increase the likelihood of them being likely benign. Then, we assessed if a subset of these VUSs could be reclassified to LP based on their enrichment in Egyptian patients (ACMG/AMP PS4 rule) or whether they reside in a previously defined mutational hotspot region that has been reported to be enriched for HCM-associated variation (modified ACMG/AMP PM1 rule) ^13,32^.

Our analysis showed that 1 VUS (*MYBPC3* c.305C>T [p.Pro102Leu]) was observed in two Egyptian controls and thus could be reclassified to likely benign according to the ACMG/AMP rules BS1 and BP4 ^29^. It is worth mentioning that here variant rarity was defined using gnomAD, which lacks sufficient data from MENA populations. Given that variant frequencies may differ across diverse ancestral populations, it is not unexpected to observe “rare” variants in gnomAD to be more prevalent in Egypt (more details in Supplementary Figure 1).

We then adopted the modified ACMG/AMP PS4 rule proposed by ClinGen, whereby variants prevalent among at least 2, 6 or 15 patients could activate the PS4-supporting, moderate and strong rules, respectively ^32^. The PS4 evidence strength levels associated with these specific patient counts were based on likelihood ratios with ideal target thresholds set by ClinGen of 10 (PS4-supporting), 30 (moderate) and 100 (strong), respectively ^32^. Of all rare variants identified in Egypt HCM patients, irrespective of variant classification, 48,3 and 1 distinct variant(s) were recurrently found in n>=2, n>=6 and n>=15 patients, respectively. Application of the modified PS4 rule upgraded the classification of 6/104 VUSs to LP (5.8%) and 10/30 LP variants to P (33.3%).

We previously identified specific regions in sarcomeric genes in which rare non-truncating variants were significantly enriched in >6,000 HCM patients of predominantly European-ancestry (LMM/OMGL) compared to controls ^21^. For variants residing in these case-enriched clusters, the etiological fraction (EF) was calculated as it provides a quantitative estimate of the probability that a rare non-truncating variant identified in an individual with HCM is disease-causing. Based on the EF findings, the authors proposed an adaptation to the ACMG/AMP guidelines whereby rare variants located in an HCM cluster with an EF≥0.95 could activate the PM1 hotspot rule at a modified strong evidence level (leading to the re-classification of VUS to LP).

As the relative case-control frequencies can differ across population groups, similar enrichment needs to be demonstrated in the population of interest before adopting these modified guidelines. Given the abundance of rare non-truncating variants observed in *MYH7* in the Egypt HCM cohort, we sought to validate the EF-based PM1 ACMG/AMP rule for *MYH7* variants (there were insufficient variants to accurately assess clustering in the troponin genes). First, we examined whether the distribution and enrichment of *MYH7* non-truncating variants was similar between the Egypt and the LMM/OMGL (n=6,112) HCM datasets. As depicted in Figure 4A, a similar distribution of rare missense *MYH7* variants was observed between Egyptian and LMM/OMGL HCM patients, with rare variants in the pre-defined HCM cluster (residues 167-931) significantly enriched in the Egypt HCM cohort over Egypt controls (13.04% vs. 0.25%, p-value=2.2×10^×16^). As the EF for Egyptian variants within the pre-defined HCM cluster was 1.0 (95%CI: 0.93-1.0), we applied the EF-based PM1_strong ACMG/AMP on rare Egyptian *MYH7* missense variants, irrespective of prior variant classification. A total of 48 missense *MYH7* variants were identified in the Egypt HCM cohort (Supplementary Table 2). Application of the EF-based PM1_strong rule upgraded the classification of 23 VUSs to LP and three LP variants to P, which increased the overall proportion of clinically actionable (i.e. P/LP) distinct *MYH7* missense variants from 35.4% to 83.3%.

**Figure 4:**
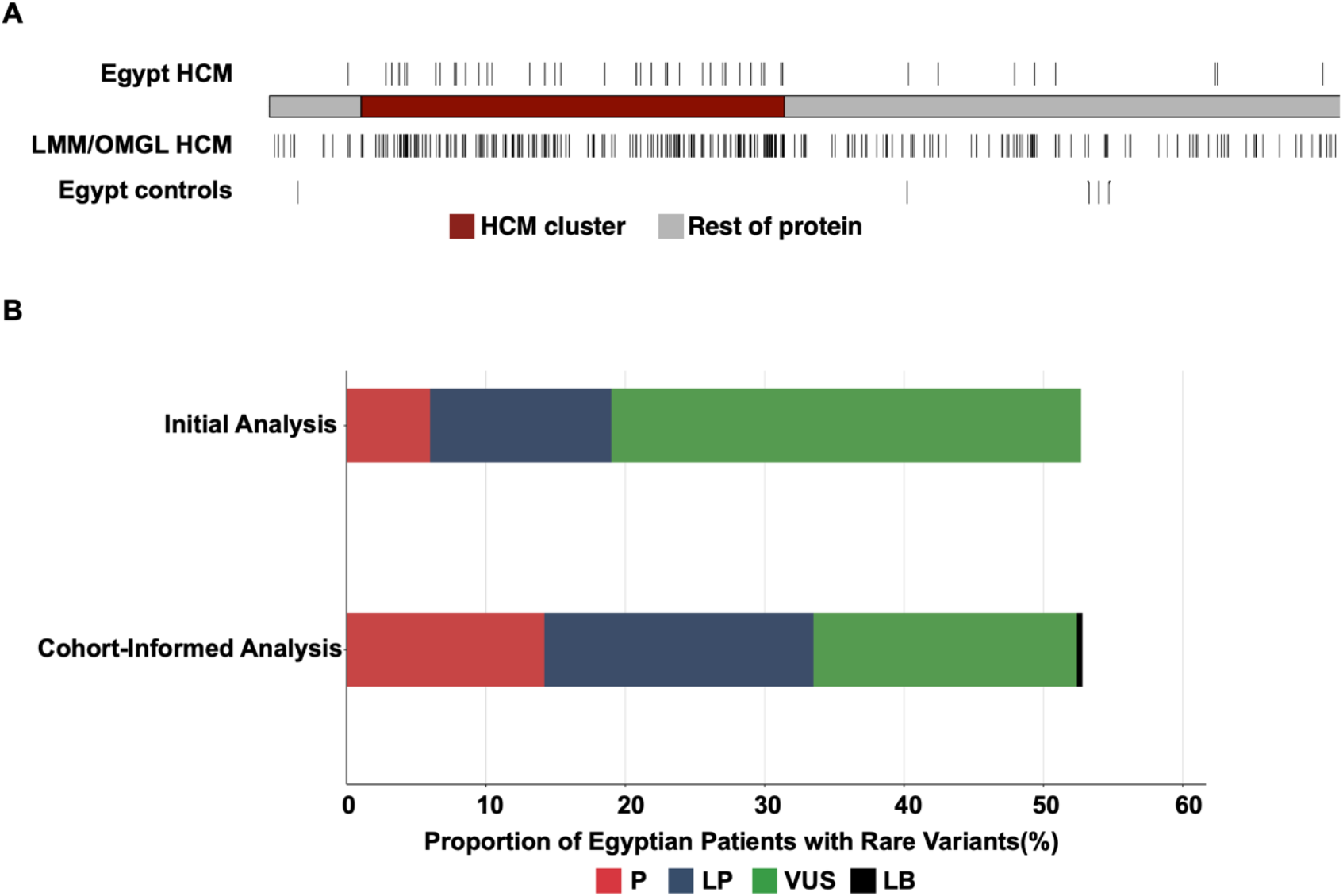
**A) Similar enrichment of rare, missense *MYH7* variants between the Egypt- and large-scale HCM cohort of predominantly European ancestry (LMM/OMGL n=6,112) in the predefined HCM cluster (red) (residues 167-931)**. Rare variants in the HCM cluster were enriched in the Egypt HCM cohort over Egypt controls with an Etiological Fraction (EF) of 1.0. **B) Application of ClinGen’s modified ACMG/AMP PS4 rule and Walsh *et al***., **2019 proposed EF-based PM1 strong rules in the Egyptian case-control cohort increases the proportion of Egypt HCM patients with clinically actionable variants from 19% to 33.4%**. Rare variants were initially classified based on the ACMG/AMP guidelines (initial analysis) and then reclassified after integrating ancestry-matched controls into the analysis (cohort-informed analysis) and adopting Egypt-specific PS4 and the modified EF-based PM1 strong rules. Cohort-informed analysis revealed that *MYBPC3* p.Pro102Leu was prevalent in the Egypt control cohort (MAF=0.0025) and thus was reclassified from VUS (in case-level analysis) to likely benign (LB, activated ACMG/AMP rules: BS1&BP4). P: pathogenic, LP: likely pathogenic, VUS: variant of uncertain significance.

Taken together, applying the above-mentioned approaches to variant interpretation increased the overall proportion of Egypt HCM patients with clinically actionable variants from 19% to 33.4% (relative increase from 40.8% to 60.4% in the patient subgroup with rare variants in validated HCM genes) (Figure 4B).

### Enrichment of non-rare variants in Egypt HCM patients

A growing body of evidence suggests that low frequency variants could contribute to disease risk with intermediate effects ^33^. These variants can be identified through burden tests showing an enrichment in patients followed by functional analyses to investigate their effect on the disease phenotype. Therefore, we sought to assess if low-frequency variants (i.e. 4×10^×5^< gnomAD FAF_popmax_< 0.01) were enriched in the Egypt HCM cohort compared to Egypt controls. A total of 20 non-rare protein-altering variants in validated HCM genes were observed in more than one patient in the Egypt HCM cohort (Supplementary Table 10). Of these, *MYBPC3* c.1321G>A [p.Glu441Lys] and *TNNT2* c.832C>T [p.Arg278Cys] were nominally enriched in Egyptian patients over controls (4.67% vs. 2%, p-value=0.03, OR=2.4 (95%CI: 1.0-6.2) and 1.17% vs. 0%, p-value=0.04, OR=10.2 (95%CI: 0.6-182.3), respectively) (although not statistically significant after multiple testing correction). While p.Arg278Cys is enriched in HCM in both Egyptian and European-ancestry cohorts, including the analyzed UK cohort, p.Glu441Lys is infrequently observed in most European HCM cohorts (Supplementary Table 11) ^34–37^. This suggests that this variant could be more relevant to Middle Eastern populations. Several publications reported the frequent co-occurrence of these variants with other sarcomeric variants suggesting that they may act as disease modifiers^33,38–41^. Notably, 37.5% (n=9/24) and 33.3% (n=2) of Egyptian patients with p.Glu441Lys and p.Arg278Cys, respectively, carried another rare sarcomeric variant, including two patients who carried p.Glu441Lys in homozygosity (Supplementary Table 11).

### Higher prevalence of *TRIM63* biallelic variants in the Egypt HCM cohort compared to Europeans

Salazar-Mendiguchía *et al*., 2020 have recently proposed that *TRIM63* is a recessive HCM gene associated with concentric LVH and a high rate of LV dysfunction following the identification of homozygous or compound heterozygous variants in 0.4% of a predominantly European (>90%) HCM cohort ^42^. We identified seven homozygous and one compound heterozygous *TRIM63* variants in the Egypt HCM cohort (total=8/374, 2.14%) (Supplementary Table 9). No biallelic *TRIM63* variants were identified in the Egypt control cohort. Similar to the recent study, c.224G>A [p.Cys75Tyr] (1.34%) and c.739C>T [p.Gln247Ter] (0.53%) (both homozygous) were the most frequently identified *TRIM63* biallelic variants in the Egypt HCM cohort ^42^. The missense variant Cys75Tyr was observed in heterozygosity in 0.5% (2/400) of the Egypt control cohort and in two individuals from the North African sub-populations from the GME study (in hetero- and homozygosity), which indicates that this variant could be particularly prevalent in North African/MENA populations ^43^. A markedly higher frequency of biallelic *TRIM63* variants was observed in the Egypt HCM cohort compared to both the analyzed UK HCM cohort and the recently published European-ancestry enriched cohort (2.14%% vs 0.16%, p-value=0.0019 and vs. 0.4%, p-value=5.8×10^×5^, respectively) ^42^.

## Discussion

This study represents the largest and most comprehensive analysis of HCM in the MENA region to date. Here, we report the results of genetic analysis of 514 unrelated Egyptian HCM patients and 400 ancestry-matched controls. We show that cohort-level analysis of rare variation in the hitherto understudied Egyptian population provides accurate interpretation of variants rarely observed in published HCM patient groups. In addition, we demonstrate that studying consanguineous populations allows for distinguishing between the relative pathogenicity and penetrance of disease-causing genes.

First, we show that the proportion of patients with rare variants in validated HCM genes was higher in the Egypt HCM cohort compared to UK HCM (52.7% vs. 32.0%). Recent data from ShaRe registry demonstrated that patients carrying sarcomere variants presented with HCM at an earlier age and had a worse prognosis ^10^. Here, Egyptian patients presented with a higher yield of sarcomeric variants compared to the UK HCM cohort (44.8% vs. 29.5%).

Patients analyzed here were admitted to AHC, which is a tertiary referral center. Thus, they typically presented with severe clinical manifestations at admission. This may explain the markedly younger age of onset (by ca. 20 years) and more severe LVH observed in the Egypt HCM cohort compared to UK HCM. To account for a possible referral bias, we compared the proportion of genotype positive patients attributable to each gene between the Egypt and UK HCM cohorts.

*MYBPC3* and *MYH7* accounted for the majority of rare variation, as with all HCM cohorts. However, some key differences are observed between the Egypt HCM and UK HCM cohorts. For example, we have recently reported the identification of a new class of pathogenic variants in HCM; a nonsense mediated decay-incompetent frameshift variant (c.5769delG) in *MYH7*, which was recurrently observed in Egyptian patients (3.3%) and absent from large-scale (n>6,000) HCM patients of predominantly European ancestry ^16,18^. Also, the Egypt HCM data shows an enrichment of *JPH2* missense variants (5.2% Egypt HCM vs. 1.4% UK HCM genotype-positive patients) which rarely cause HCM outside of founder populations. This is largely explained by two recurrent variants that have not been previously reported ^25^. One of these variants, p.Thr161Ala, occurs at the same position as another pathogenic variant (p.Thr161Lys) recently reported to be a founder in the Finnish population ^44^. This may denote a particular pathogenic hotspot in *JPH2* although further segregation and/or functional data are required to enable a more accurate classification of these variants.

The main unique finding, however, relates to the clear prevalence of HCM minor genes, particularly *MYL2, MYL3* (MLC) and *CSRP3*, in the Egyptian population and the enrichment of homozygous variants in these genes. A noticeably higher proportion of rare variants in MLC genes was observed in the genotype-positive Egypt HCM cohort compared to UK HCM (10.33% vs. 2.28%), including five recurrent variants that were absent from the latter cohort. Interestingly, a substantial proportion of MLC variants (25.6%) were observed in the homozygous state, suggesting such variants are less penetrant in heterozygosity. In agreement with this notion, a study by Claes *et al*., 2016 demonstrated that the expression of the Dutch founder *MYL2* missense variant p.Glu22Lys in isolation was associated with low HCM penetrance and a benign disease manifestation and that the presence of an additional risk factor, including hypertension, obesity or the co-occurrence of another sarcomeric variant, increased the disease penetrance from 36% to 89% in *MYL2*-variant carriers ^45^. In addition, the authors reported that patients carrying p.Glu22Lys in homozygosity were diagnosed with end-stage heart failure due to HCM in their 30s ^45^. It is therefore plausible to conclude that the high consanguinity rates observed in the Egypt HCM cohort enabled MLC variants to occur in a more penetrant homozygous state. This may explain, at least in part, the relatively higher proportion of rare MLC variants observed in the analyzed consanguineous Egypt HCM cohort compared to UK HCM. While biallelic truncating variants in *MYL2/MYL3* are associated with severe early onset cardioskeletal myopathies, only two homozygous missense variants in *MYL3* have previously been reported - p.Glu143Lys^46,47^ and p.Ala57Asp^48,49^. It is noteworthy that the homozygous expression of the non-rare variant p.Ala57Asp, which was observed in HCM patients of Iranian and Tunisian origins born to consanguineous parents^48,49^, was also observed in the Egypt HCM cohort (in two homozygous and one heterozygous carriers). The homozygous patients presented with HCM at a relatively younger age and had a greater LVWMT (27+2.8mm vs. 16mm) compared to the heterozygous carrier. Although a previous functional study with iPSC-CMs did not demonstrate a phenotype for this variant in either the heterozygous or homozygous state^50^, the fact that is has now been detected as a homozygous variant in four HCM cases (and not detected in our healthy control cohort) suggests it is contributing to the phenotype.

Similar to MLC-encoding genes, variants in *CSRP3* were more likely to present in homozygosity (33.3%) than the major sarcomere-encoding HCM genes (*MYH7, MYBPC3* and thin filament genes, 5-7%). Two of the three homozygous *CSRP3* variants were truncating (c.415-1G>C and c.414+1G>T). These variants were observed in one and three Egyptian patients, respectively. Patients carrying these variants presented with HCM at an average age of 45.5±8.7 and a LVMWT of 16.5±2.4mm. It is noteworthy that c.414+1G>T was also observed in heterozygosity in one Egyptian control and HCM patient, respectively. The control did not carry any other rare HCM-associated variant whereas the HCM patient carried the pathogenic *MYH7* c.5769delG variant (and the *JPH2* p.Ser99Asn VUS), suggesting that in the heterozygous state this variant requires an additional mutational burden to express the HCM phenotype. To date, only three knockout *CSRP3* HCM patients have been reported in Poland and France, two of whom were confirmed to be born to consanguineous parents^51,52^. These patients were diagnosed with adult-onset HCM and one patient underwent Implantable cardioverter defibrillator implantation in her 30s ^51,52^. Together, these studies suggest that full *CSRP3* gene knockouts, in contrast to the severe early-onset phenotypes observed with biallelic truncating variants in genes like *MYBPC3* and *ALPK3*, are associated with more typical adult-onset HCM, despite the fact that most reports for *CSRP3*-associated HCM involve heterozygous variants and dominant inheritance.

Taken together, these findings demonstrate that this study is uniquely able to examine the frequency of homozygous variants in dominant HCM genes using a large consanguineous cohort to inform the relative penetrance of variants in these genes. The recurrence of these variants in the Egyptian cohort, which may indeed have generally higher prevalence in other consanguineous populations of the MENA region, exemplifies the need to integrate new guidelines tailored to the classification of homozygous variants into the current ACMG framework in order to enhance the utility of clinical genetic testing in other consanguineous populations.

The observed high prevalence of consanguinity (28%) in the Egyptian cohort analyzed here has also enabled us to validate recent findings reporting the association of the recessive gene *TRIM63* to HCM ^42^. Indeed, the prevalence of biallelic *TRIM63* variants was markedly higher in Egyptian patients compared to HCM patients of European ancestries suggesting that recessive inheritance patterns of HCM are more likely to occur in consanguineous populations of the MENA region.

Our data also highlight the value of performing ancestry-matched case-control analysis coupled with standardized guidelines and quantitative approaches to variant interpretation to accurately classify variants observed in underrepresented populations ^13,29,32^. We demonstrate that these approaches enabled 23.1% of distinct VUSs identified in the Egypt HCM cohort to progress to likely pathogenic, enabling a more equitable application of clinical genetic testing in the region. Expanding the MENA dataset in resources like gnomAD will further improve the interpretation of VUSs by providing more accurate frequencies of these variants in the population.

It is noteworthy that despite the relatively severe phenotypes observed in our cohort, almost 50% of HCM patients in this study presented with an unexplained genetic etiology. This finding is not unexpected as recent evidence suggests that the genetic etiology of HCM is more complex and that common variants with individually small (but collectively large) effect sizes could explain some of the missing heritability of the disease ^53,54^. This study is not designed to assess common variants or validate recently described European-ancestry HCM polygenic risk scores (PRS)^54^. However, a recent HCM genome-wide association study (GWAS) identified a common *BAG3* missense variant p.Cys151Arg, as one of the two most strongly associated loci for HCM and which is covered by the targeted gene panel used in this analysis^54^. The OR of p.Cys151Arg was highly similar between the Egyptian cohorts and the published European datasets, 1.47 (95%CI: 1.12-1.95) and 1.42 (95%CI: 1.29-1.55) respectively ^54^, although the population frequency for the minor risk allele is lower in Egypt (control MAF=0.12) than Europe (gnomAD-non-Finnish European MAF=0.22). This suggests that there will be likely many shared loci between populations, although well-powered GWAS in MENA populations are warranted to accurately quantify HCM PRS for future clinical use.

Low frequency variants with effect sizes that are intermediate between common GWAS-derived and rare Mendelian variants are now also emerging as additional genetic risk factors. For example, a recent study by Pua *et al*. identified two low frequency troponin variants (*TNNI3* p.Arg79Cys OR=3.3 (96%CI: 1.5-7.2), MAF=0.018 and *TNNT2* p.Arg286His OR=14.1 (95%CI: 6.0-33.0), MAF=0.022) that are associated with HCM in patients of Chinese ancestry ^55^. Similarly, we observed two low frequency variants *MYBPC3* p.Glu441Lys (OR=2.4 (95%CI: 1.0-6.2), MAF=0.025) and *TNNT2* p.Arg278Cys (OR= 10.2 (95%CI: 0.6-182.3), MAF=0.006) that were nominally enriched in the Egyptian HCM cohort. While the *TNNT2* p.Arg278Cys variant is also consistently enriched in European ancestry cohorts, the *MYBPC3* variant p.Glu441Lys is clearly more prevalent in the Egypt HCM cohort, suggesting that it could be more relevant to Egyptian patients ^34–37^. In agreement with literature findings, both variants were frequently observed in conjunction with another sarcomeric variant in Egyptian patients, which suggests their modifier effect on the phenotype ^38–41^. However, functional studies will be required to definitively ascertain their effect on HCM phenotype.

A limitation of the study is that the targeted gene panel used does not include genes that have been recently shown to be associated with HCM (*ALPK3, FHOD3* and *FLNC* ^56–59^). In addition, the study design did not account for common and copy number variations, non-coding variants or epigenetic factors, which may contribute to HCM pathogenesis. Finally, larger control datasets are needed to fully characterize rare variation in MENA populations.

In conclusion, we have demonstrated that detailed analysis of rare genetic variation in validated HCM genes in underrepresented populations reveals novel findings with relevance to both clinical genetic testing and our understanding of the genetic etiology of HCM. We report the contribution of minor HCM genes, rarely observed in outbred populations, to disease pathogenicity. We also show a higher prevalence of biallelic variants in the recessive HCM gene *TRIM63* in Egyptian patients suggesting that recessive inheritance of HCM is more likely to occur in consanguineous populations. These unique genetic features exemplify the value of studying diverse ancestral populations. The highly consanguineous cohort analyzed here opens new research avenues for the discovery of novel recessive genes in future exome or genome sequencing studies.

## Data Availability

All data produced in the present work are contained in the manuscript

## Acknowledgements

We would like to acknowledge Dr. Rafik Tadros and Dr. Paloma Jordà for providing us with HCM and control frequency data regarding the recently reported *BAG3* variant p.Cys151Arg. We would like to thank the AHC nursing staff and team members who contributed to the biobanking efforts in frame of the National HCM Registry.

## Web resources

OMIM, https://www.omim.org

