## Supplementary Methods for "Influence of ethnicity and consanguinity on the genetic architecture of Hypertrophic Cardiomyopathy: insights from an understudied population"

### Rare variant filtering and classification

Rare variants in validated HCM genes were defined as having a filtering allele frequency (FAF) of ≤ $4x10^{-5}$ in gnomAD based on the statistical framework proposed by Whiffin *et al.,* 2017 [18]. In this study, WES data was downloaded from the gnomAD database (https://gnomad.broadinstitute.org; version v2.1.1) and only variants with a PASS filter were included in the analysis. Variants in the sarcomere genes (*ACTC1*, *MYBPC3*, *MYH7*, *MYL2*, *MYL3*, *TNNC1*, *TNNI3*, *TNNT2* and *TPM1*) and the minor HCM genes (*PLN* and *CSRP3*) were classified into Pathogenic (P), Likely Pathogenic (LP) and Uncertain Significance (VUS) using CardioClassifier, which integrates the ACMG/AMP guidelines to aid in the interpretation of variants identified in a gene associated with an inherited cardiac condition [1,2]. The following rules are automatically activated by CardioClassifier:

- **PM1:** Mutational hotspot or well-studied functional domain without benign variation. This rule was applied at moderate level to *MYH7* missense variants located in the mutational hotspot (residues 181-937).
- **PM2:** Low frequency in population databases (i.e., FAF in ExAC < $4×10^{-5}$). In this analysis, gnomAD was used as the reference population, so gnomAD data (https://gnomad.broadinstitute.org; version v2.1.1) was downloaded for the validated HCM genes. Then, the gnomAD $FAF_{popmax}$ was then calculated using the R script "frequencyFilter" available in github (https://github.com/ImperialCardioGenetics/frequencyFilter/blob/master/src/precompute_exac_af_filter.R).
- **PVS1:** Truncating variants in genes *CSRP3*, *FHL1*, *MYBPC3*, *TNNI3*, *TNNT2* and *PLN*, which are statistically enriched in LMM/OMGL patients over controls.

- **PS4:** Variant is statistically enriched in LMM/OMGL patients over controls with the rule activated if the patient count was > 2 and the Fisher's exact test p-value was $< 1.79 \times 10^{-6}$ (Bonferroni correction).
- **PM4:** Protein length changing variants as a result of inframe insertion/deletions or stop lost.
- **PP3:** Missense variant with multiple lines of computational evidence supporting a deleterious effect. The computational prediction algorithms include: SIFT, PolyPhen2 var., LRT, Mutation Taster, Mutation Assessor, FATHMM, CADD and Grantham scores. This rule is activated if at least 5/8 tools predict a deleterious effect, with only 1 tool predicting benign and <3 with "unknown" classifications or if >3 tools have unknown variant classifications while all other tools predict a deleterious effect.
- **PM5/PS1:** Novel missense variant where a different missense variant at the same amino acid residue is classified as pathogenic or novel missense variants with the same amino acid change as an established pathogenic variant (PS1). Novel variants are defined as "pathogenic" if multiple submitters in ClinVar confer pathogenicity with no conflicting evidence.

*ACTN2* and *JPH2* are not included in CardioClassifier and thus were manually classified according to the ACMG/AMP guidelines.

Then, rare variants, which activate the PM2 rule, were manually curated using ClinVar and PubMed resources for evidence regarding the following ACMG/AMP rules:

- **PP1:** Co-segregation with disease (supporting ≥ 3 meiosis, moderate ≥ 5, strong ≥ 7)
- **PM2/PM6:** *De novo* inheritance (with/without confirmed maternity and paternity)
- **PS3:** rule relating to evidence of deleterious effects of variants was not applied as functional approaches are not well-validated for HCM variants [3]

1. Richards, S. *et al.* Standards and guidelines for the interpretation of sequence variants: a joint consensus recommendation of the American College of Medical Genetics and Genomics and the Association for Molecular Pathology. *Genet. Med.* **17**, 405–423 (2015).
2. Whiffin, N. *et al.* CardioClassifier: disease- and gene-specific computational decision support for clinical genome interpretation. *Genet. Med.* **20**, 1246–1254 (2018).
3. Walsh, R. *et al.* Quantitative approaches to variant classification increase the yield and precision of genetic testing in Mendelian diseases : the case of hypertrophic cardiomyopathy. *Genome Med.* **11**, 1–18 (2019).
