## Supplementary Data for "Influence of ethnicity and consanguinity on the genetic architecture of Hypertrophic Cardiomyopathy: insights from an understudied population"

**“Rare” variants defined using gnomAD introduced bias in comparing HCM genetic architecture between Egypt and UK HCM cases**

The definition of variant rarity in the Egyptian HCM cohort was based on 5 gnomAD populations: African/African American, Admixed American, East Asian, Non-Finnish European and South Asian while data from the MENA region was lacking in the dataset. Thus, variants considered “rare” in Egyptian cases may actually be common/non-rare in the general Egyptian population. To evaluate the bias in defining variant rarity based on gnomAD, we compared the burden of rare synonymous variants in all ICC genes between the Egyptian (i.e. Egyptian HCM cases + Egyptian controls) and predominantly European (UK cases + UK controls) cohorts. We selected synonymous variants for this analysis as they are not expected to be disease-causing and thus provide an unbiased approach to analyse inflation. We evaluated burden testing data for inflation with a quantile-quantile (Q-Q) plot and then estimated genomic inflation factor (GIF) as per the formula proposed by Guo *et al.* 2018[1].

As depicted in Supplementary Supplementary Figure **1**, the resulting distribution of the ICC single-gene p-values was characterised by a GIF of 2.770. This finding reflects an inflation of the expected- (under uniform distribution) and observed p-values (from burden testing data) above the expected GIF = 1, suggesting the presence of bias.

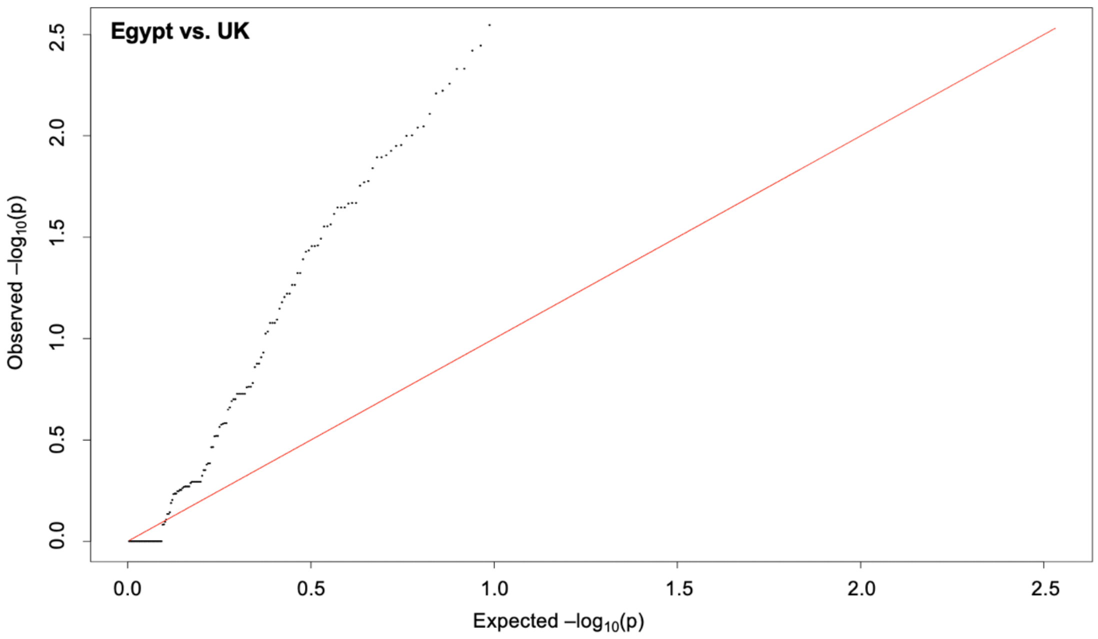


**Supplementary Figure 1: Quantile-quantile (Q-Q) plot of Egyptian vs. *UK* burden testing data based on synonymous variants.** Expected -$\log_{10}$ p-values for rare (gnomAD $FAF_{popmax} \leq 4x10^{-5}$) synonymous variants, under the uniform distribution (red line), were plotted against the observed -$\log_{10}$ p-values from the burden testing data for each ICC gene. The plot shows a discordance between expected and observed values with a genomic inflation factor (GIF) of 2.770.

The observed high GIF confirmed that the biased definition of variant rarity based on gnomAD confounded the comparison of rare variation between Egyptian and UK HCM cohorts. This bias might explain the observed higher frequency of rare variants in HCM genes in Egyptian HCM cases compared to UK (refer to Figure 1 in Manuscript).

1. Guo, M. H., Plummer, L., Chan, Y. M., Hirschhorn, J. N. & Lippincott, M. F. Burden Testing of Rare Variants Identified through Exome Sequencing via Publicly Available Control Data. *Am. J. Hum. Genet.* **103**, 522–534 (2018).

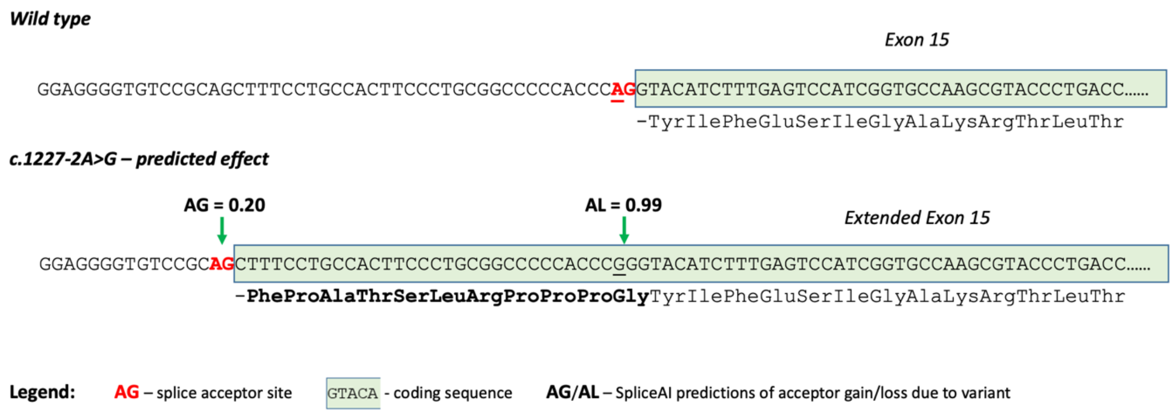


**Supplementary Figure 2: Schematic presentation of the predicted effect (by SpliceAI) of the splice acceptor variant c.1227-2A>G on the exonic sequence of M*YBPC3*.** The variant is predicted to result in an exon extension of 33 intronic bases into the *MYBPC3* transcript suggesting that it maintains the open reading frame and thus may function as an inframe insertion.
